# Empirical evidence of transmission over a school-household network for SARS-CoV-2; exploration of transmission pairs stratified by primary and secondary school

**DOI:** 10.1101/2022.02.12.22270851

**Authors:** Senna C.J.L. van Iersel, Jantien A. Backer, Rolina D. van Gaalen, Stijn P. Andeweg, James D. Munday, Jacco Wallinga, Albert Jan van Hoek, the RIVM COVID-19 epidemiology and surveillance group

## Abstract

**Background:** Children play a key role in the transmission of many infectious diseases. They have many of their close social encounters at home or at school. We hypothesized that most of the transmission of respiratory infections among children occur in these two settings and that transmission patterns can be predicted by a bipartite network of schools and households.

**Aim and methods:** To confirm transmission over a school-household network, SARS-CoV-2 transmission pairs in children aged 4-17 years were analyzed by study year and primary/secondary school. Cases with symptom onset between the 1^st^ of March 2021 and the 4^th^ of April 2021 identified by source and contact-tracing in the Netherlands were included. In this period, primary schools were open and secondary school students attended class at least once per week. Within pairs, spatial distance between the postcodes was calculated as the Euclidean distance.

**Results:** A total of 4,059 transmission pairs were identified; 51.9% between primary schoolers; 19.6% between primary and secondary schoolers; 28.5% between secondary schoolers. Most (68.5%) of the transmission for children in the same study year occurred at school. In contrast, most of the transmission of children from different study years (64.3%) and most primary-secondary transmission (81.7%) occurred at home. The average spatial distance between infections was 1.2km (median 0.4) for primary school pairs, 1.6km (median 0) for primary-secondary school pairs and 4.1km (median 1.2) for secondary school pairs.

**Conclusion:** The results provide evidence of transmission on a bipartite school-household network. Schools play an important role in transmission within study years, and households play an important role in transmission between study years and between primary and secondary schools. Spatial distance between infections in a transmission pair reflects the smaller school catchment area of primary schools versus secondary schools. Many of these observed patterns likely hold for other respiratory pathogens.

## Background

Children play a key role in the transmission of many infectious diseases, and receive most vaccines. Detailed understanding of which children interact with each other (age, sex, location, social group, vaccination status, etc), and which of these interactions lead to transmission is critical to define and implement interventions to curtail transmission, or to assess the risk for outbreaks when vaccination rates decline. As the public health response implementing interventions or vaccination campaigns is often organized by local authorities, this understanding should ideally be available on a local level.

Contact surveys show that primary school children have most of their contact time at school or at home (1). We hypothesize that transmission among children can be described as a bipartite network of schools and households as nodes, with children as edges, or similarly as a network of schools as nodes with household links as edges (2). Such a description could provide a framework to evaluate the impact of interventions on the transmission at both a national and a local level, while accounting for variation in school level (primary, secondary) and school size. Spatial spread of infection can be included with the location of schools and their catchment area. We have shown that it is possible to construct such a network for England using governmental administrative data (2).

To evaluate if transmission among children indeed follows the bipartite structure of schools and households, we explored the reported setting of SARS-CoV-2 transmission (school, home, and somewhere else) in transmission pairs of school-aged children during the SARS-CoV-2 pandemic in the Netherlands.

## Methods

We selected over 4,000 reported transmission pairs between children aged 4-17 within the national infectious disease notification registry with symptom onset between the 1^st^ of March 2021 until 28^th^ of March 2021 for infectors and between the 1^st^ of March 2021 and the 4^th^ of April 2021 for infectees.

During this period, primary schools were open full-time for in-class education and secondary schools were offering in-class education at least one day per week for each student with physical distancing and mask wearing within the secondary school. Secondary schools remained open for exam students and students in a vulnerable position. Primary and secondary school children could attend school only when free of COVID-like symptoms or by showing a negative SARS-CoV-2 test result. School contacts of test-positive cases were quarantined. After-school childcare was closed for children whose parents were not essential workers, and most other group-based activities for children of all ages were not allowed. Free SARS-CoV-2 testing was available for everybody with COVID-like symptoms, with close contact with test-positive cases, and who had travelled abroad. Over-the-counter self-swab SARS-CoV-2 antigen tests were not available until March 30, 2021. Selection of cases was based on self-reported date of symptom onset, or when this date was not available, the symptom onset date was imputed with the day of the positive test minus the median number of days between onset and a positive test, or date of registration minus the median number of days between onset and registration.

A transmission pair consists of two laboratory-confirmed (PCR or antigen test) SARS-CoV-2 cases, with an epidemiological link that suggests direct transmission as identified through case finding and contact tracing by the Municipal Health Service. Reciprocal pairs and triangular chains of cases are excluded from our analysis since it is not possible to uniquely identify the infector and infectee in the pair. The registered settings of transmission are categorized as ‘home’, ‘school’, and ‘other’.

We estimate the school year using the age of the child on the 1^st^ of October 2020, as age minus three years. This estimation has an approximated accuracy of 80% in estimating the actual school year [Personal communication the Education Executive Agency of the Dutch Ministry of Education, Culture and Science (DUO)]. School years 1-8 were assigned to primary school and are labelled as P1,P2,…,P8; school years 9-14 to secondary school, S1,…,S6.

To explore the transmission between school years, we looked at the absolute difference in transmission between all pairs of different school years. For each school year combination, we looked at the number of transmission pairs from one school year to the other and vice versa and obtained the absolute difference in the number of pairs. As sometimes the difference was small, or there were a small number of pairs in total, we only considered the difference informative when the probability that this difference would occur as sampled from a binomial distribution (with p = 50%) was less than 5%.

The spatial distance between two children in a pair was obtained as Euclidean distance using their geo-mapped full residential postcodes. If transmission occurs in schools, we expect these distances to be smaller for primary schoolers than secondary schoolers as the catchment area of primary schools is smaller than that of secondary schools.

To explore the transmission between school and household via siblings, we selected all reported transmission triplets of children aged 4-17 where the first transmission had setting ‘school’ and the second transmission setting ‘home’ or vice versa, and where at least one of the three children involved had a different postcode to exclude triples in the same household.

## Results

In the period 1^st^ of March 2021 until 28^th^ of March 2021, there were 175,169 test-positive cases in the Netherlands, with 44,132 (25.2%) unique cases identified as infector. These infectors account for 63,581 transmission pairs with infectee symptom onset until the 4^th^ of April (Table 1). Most of these pairs consisted of an adult as infector and either an adult or a child too young to attend primary school as infectee (44,590), 14,484 consisted of one child of school age and one other person not of school age, and 4,059 pairs (6.3% of the total number of pairs) consisted of two children of school age. The rest of the pairs (448) had a child too young to attend primary school as infector and either an adult or child too young to attend school as infectee. Of the 31,909 test-positive school aged children between the 1^st^ of March 2021 and 28^th^ of March 2021, 18,260 (57.2%) attended primary school and 13,649 (42.8%) attended secondary school. The percentages were similar for the infectors in the pairs, with 2,607 (64.2%) primary school infectors and 1,452 (35.8%) secondary school infectors. There were 6,668 unique school aged children identified as infector (20.9%), of which 3,229 infected another school aged child. Symptom onset was known for 84.7% of the infectors and 67.0% of the infectees, the rest was imputed. Full 6-digit postcode information was available for 99.7% of all cases in pairs consisting of school-aged children, and the possible setting of infection was reported for 99.5% of the infectees.

**Table 1 –.**
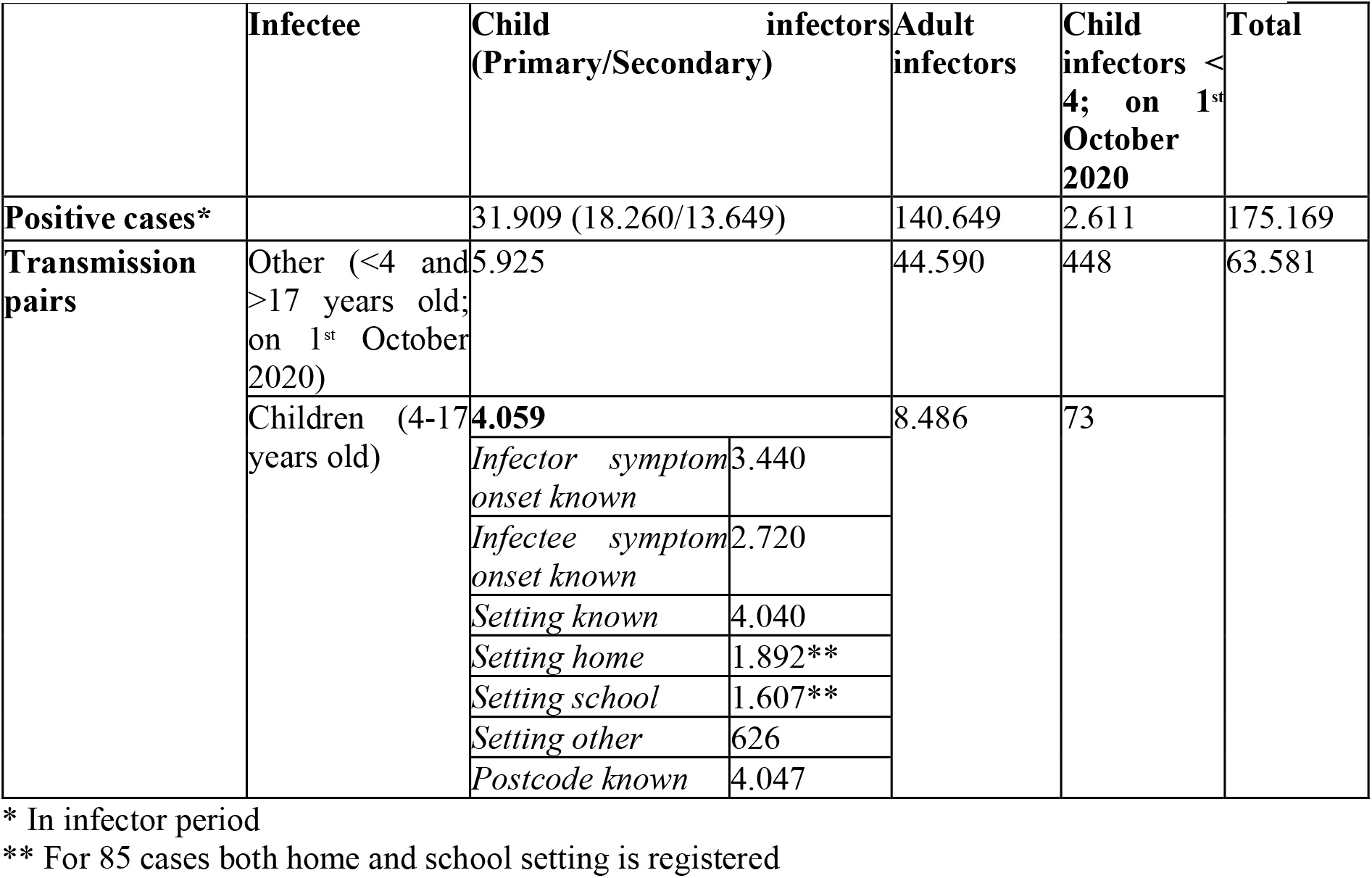
Descriptive statistics of positive SARS-CoV-2 cases and transmission pairs

Most of the selected transmission pairs consisted of children in the same school year, and most pairs were among the oldest school years of primary and secondary school (Figure 1). There is a distinct pattern of transmission per setting, where transmission within the same year happens at school, and transmission between school years happens at home. This is especially so for children in primary school, as 86.0% of the transmission pairs consisting of children in the same school year occurred at school. Respectively 65.4% and 62.4% of the transmission pairs of primary and secondary school infectors with infectees in a different school year, thus including primary-secondary school pairs, happened at home. Children in secondary school within the same school year get infected more often in other settings than school and home (52.0%), particularly those in class S5 and S6.

**Figure 1 -.**
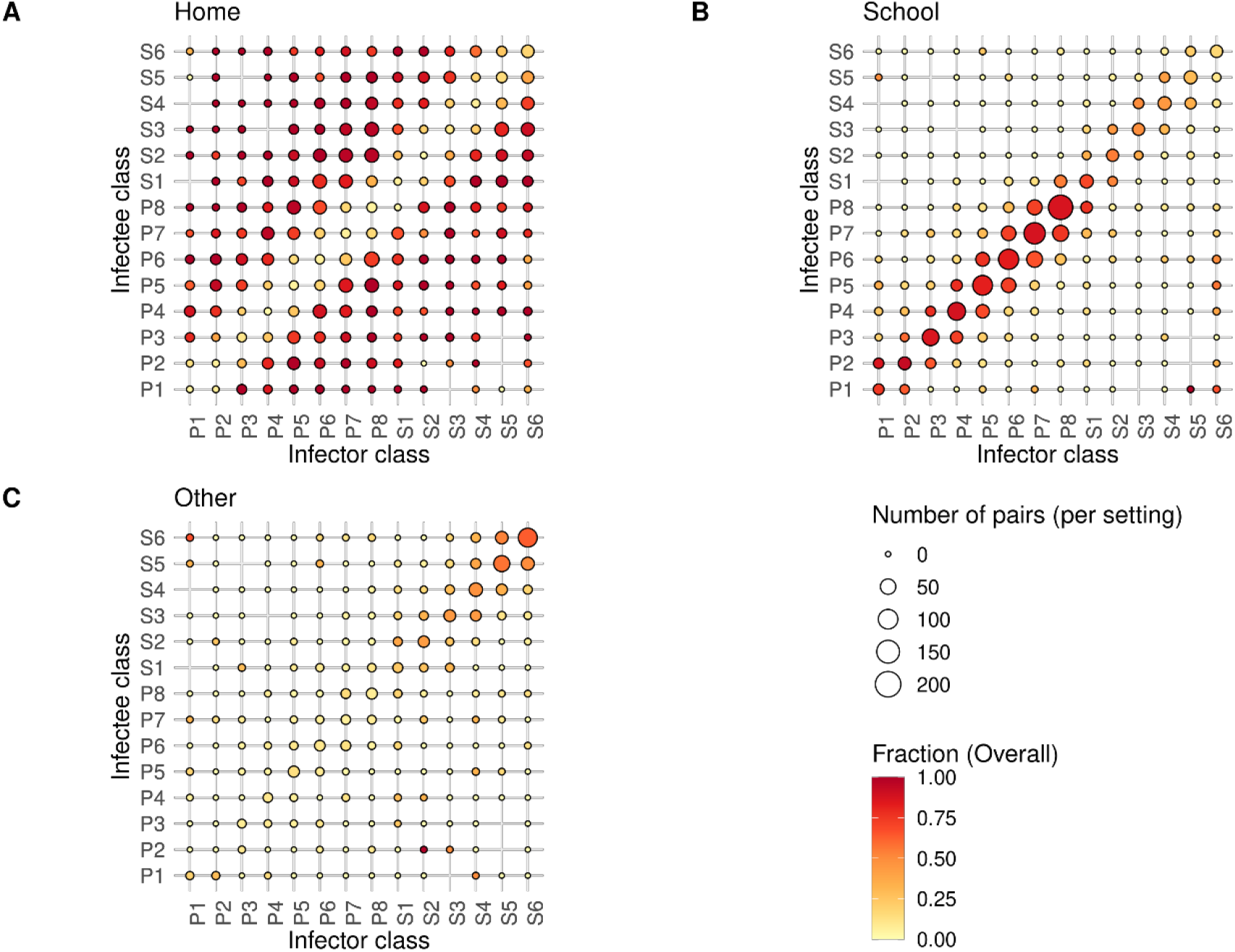
Transmission pairs by class (primary school P1-P8, secondary school S1-S6) in the period between the 1^st^ of March and the 28^th^ of March for infectors and between the 1^st^ of March and the 4^th^ of April for infectees. The size of the bubbles shows the number of pairs per setting. Transmission pairs with an unknown setting are excluded. The color of the bubbles indicates the fraction overall of transmission in **A -** the home setting. **B -** the school setting. **C -** other settings than household and school setting.

Children in higher classes of primary (P5-P8) and secondary (S5-S6) school were more often the infector than the infectee compared to children in lower classes (Figure 2). Children in higher classes of primary school were also more often the infector of secondary school students than the other way around (Figure 2).

**Figure 2 –.**
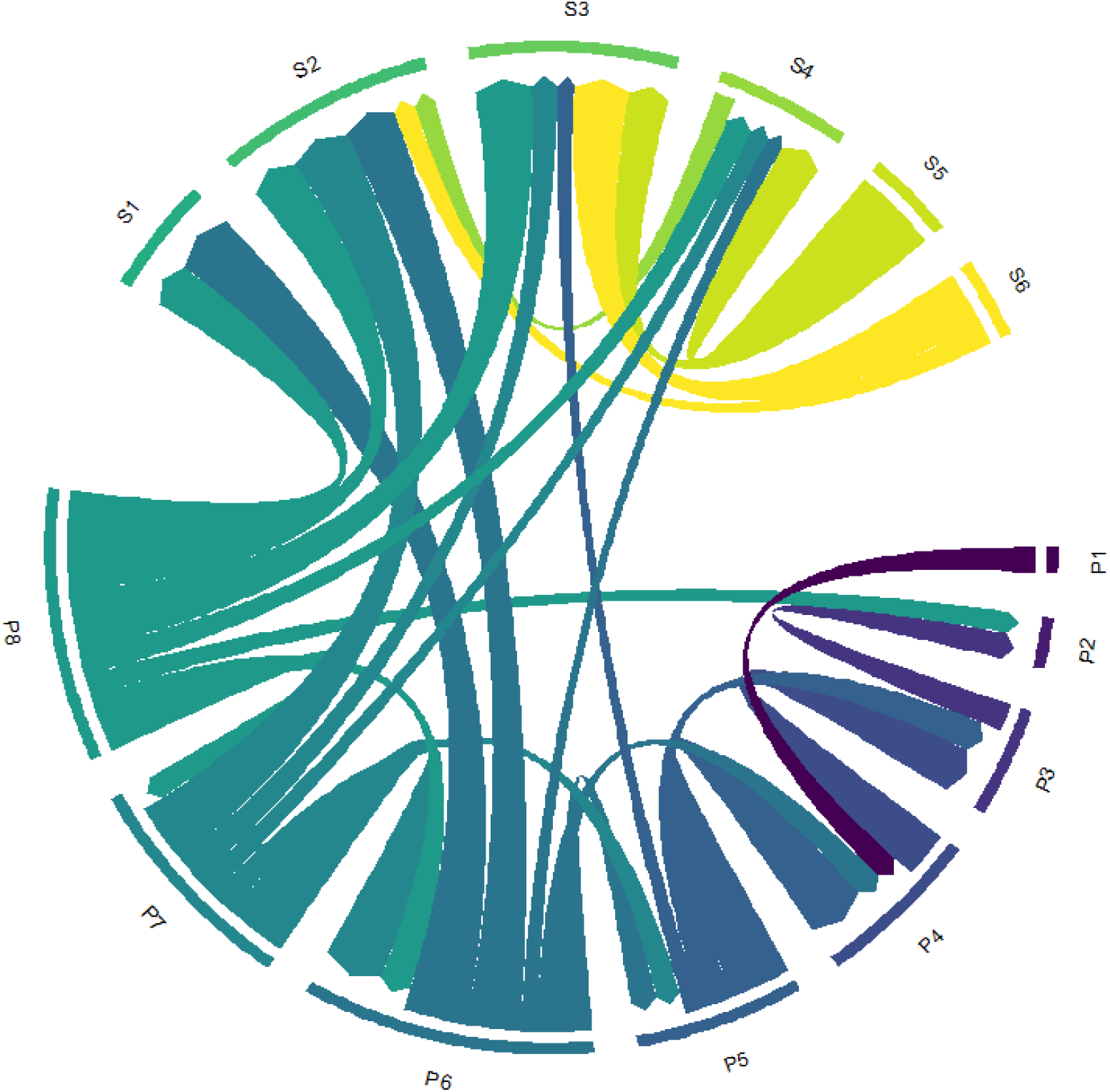
Chord-diagram showing the absolute difference in transmission pairs between school years (difference A→B, A←B). Only those arrows are shown for which there is a significant asymmetry in the direction of pairs. Colors indicate the infector age class.

The distance between cases in pairs of primary schoolers is on average 1.2 km, increasing up to 4.1 km for cases in pairs of secondary schoolers (Figure 3). The same pattern holds when excluding pairs that share the same postcode, thus excluding children in the same household. The average distance between cases in pairs of primary schoolers with a different postcode is 1.9 km. For pairs of secondary schoolers the average distance between cases is 6.1 km.

**Figure 3 –.**
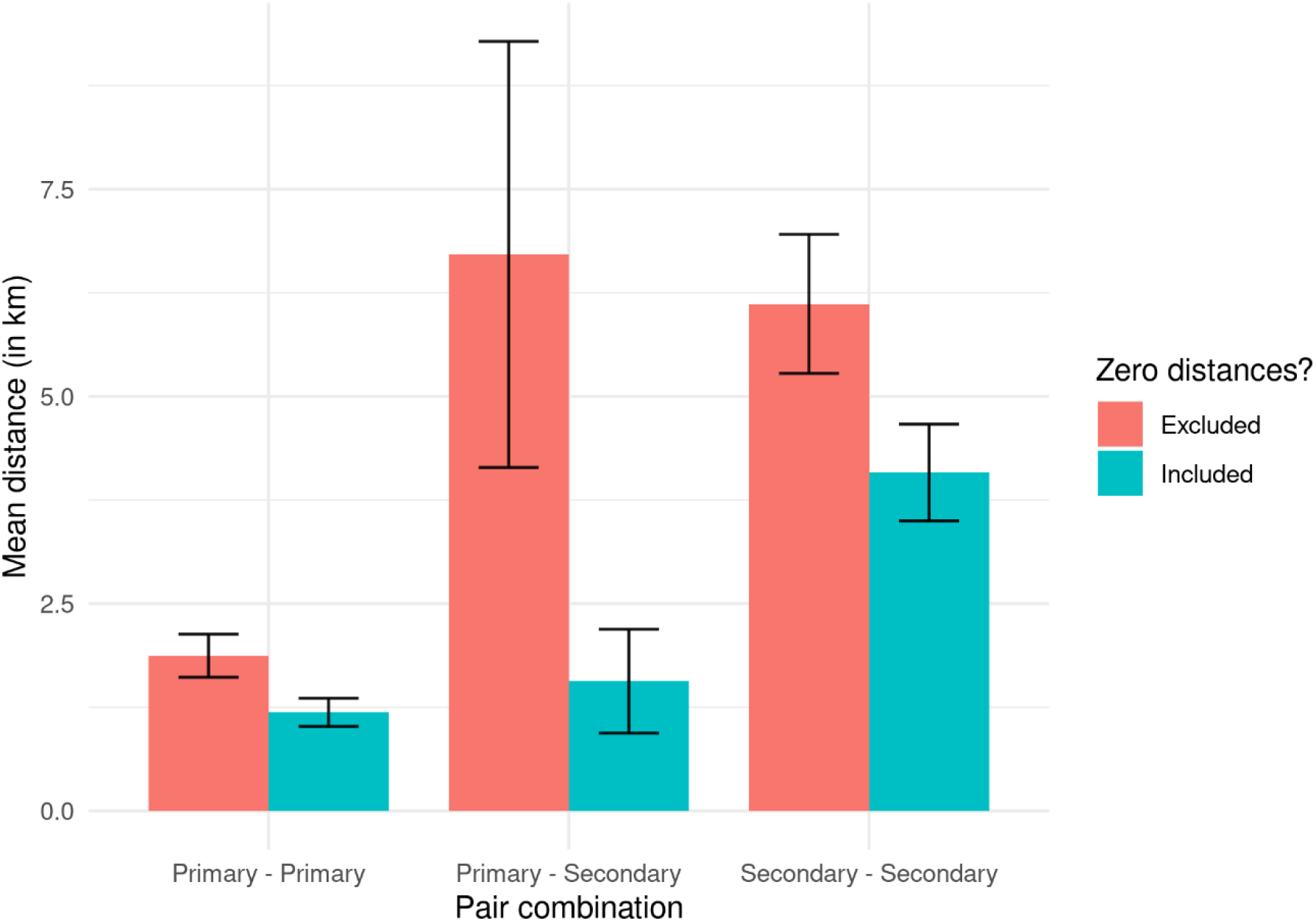
Mean distances in kilometers for primary and secondary school transmission pair combinations with confidence interval. Red bars indicate mean distances without transmission pairs within the same PC6 (zero distance), blue bars indicate mean distances including transmission pairs within the same PC6.

In around one third of the identified transmission chains of three children there is transmission between children of primary school age and children of secondary school age (Figure 4). Also, children more frequently get infected at school and subsequently infect a third person at home than vice versa. Over the study period where primary schools were open and secondary schools were partially open, primary school children make up a larger share in the transmission chains than secondary school children.

**Figure 4 –.**
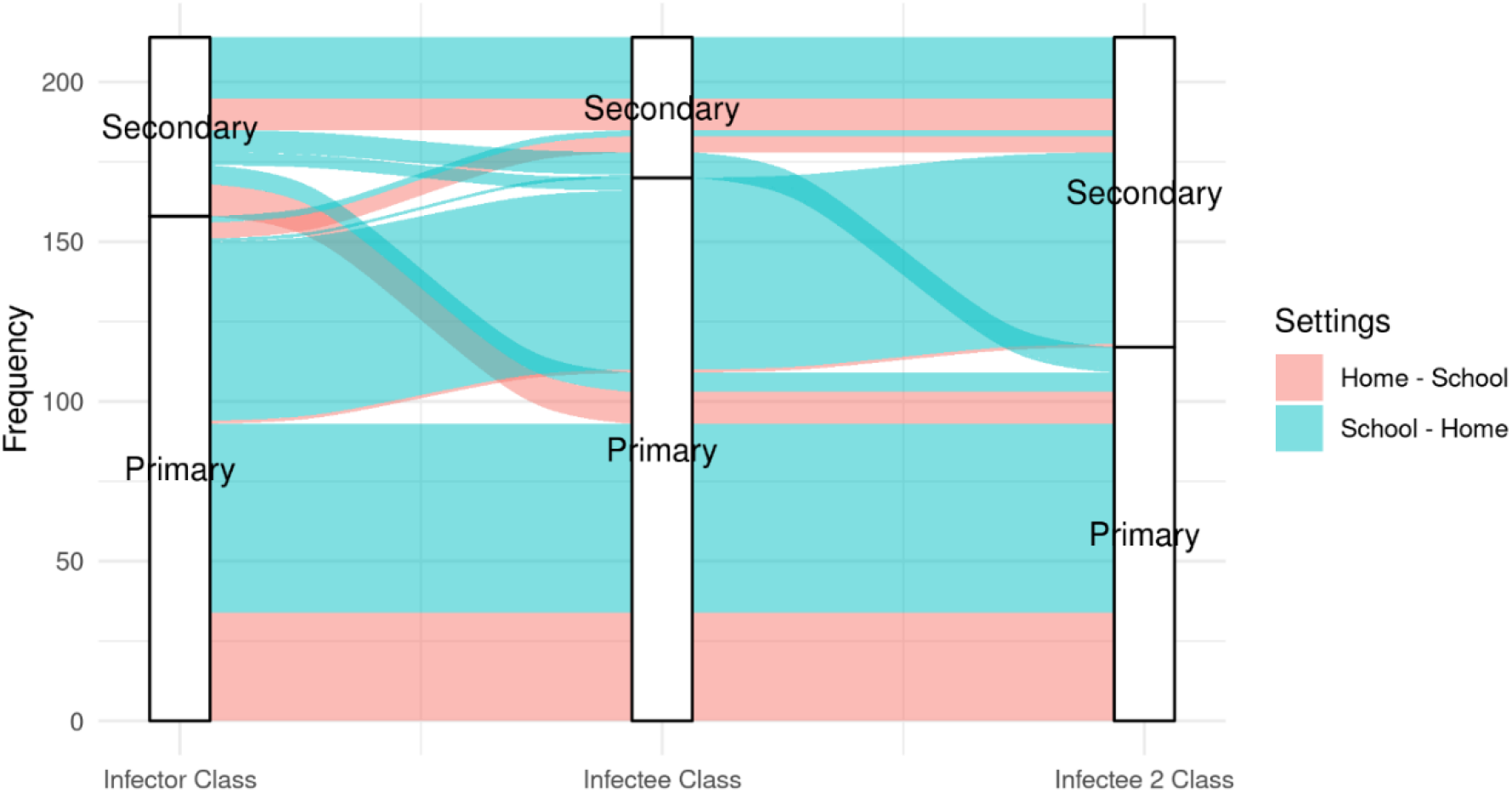
Transmission chains with frequency of infections between primary and secondary school, with at least one infected child not living in the same household as the others. Only chains are shown where the setting of transmission of the first pair in the chain is either ‘home’ or ‘school’, with the setting of transmission for the second pair in the chain being the other of the two settings. Red indicates that the transmission of the first pair took place in the ‘home’ setting and thus the transmission setting of the second pair is ‘school’. For blue, it is the other way around.

## Conclusion and discussion

Our results show that SARS-CoV-2 transmission between primary school children in the Netherlands follow a bipartite network between school and households during the pandemic. We observe that transmission within the same (school) year happens at school and transmission between (school) years at home, and show that siblings link different schools in transmission (primary and secondary). Furthermore, we show that the spatial spread of infections is further apart in secondary school pairs compared to primary school pairs, in line with the wider catchment areas of secondary schools. Our results provide an empirical justification to use of a school-household network to understand transmission patterns among primary school children for the pandemic period. We also show that in this period there were distinct roles for the highest years in primary school and lowest years in secondary school, highlighting that it is important to understand interventions in primary and secondary schools, as well as susceptibility and infectiousness by age.

Although our observations are about SARS-CoV-2 in a pandemic situation, we would argue that our observations are informative for other respiratory infections such as influenza and for the non-pandemic period. Our observations highlight a transmission pattern that follow school years, instead of random mixing within the school, and assign an important role for households connecting school years. This is in line with a detailed contact study in primary school (3), pre-pandemic self-reported time-use data in the US (1), as well as in a previous outbreak investigation in the US (4), and a recent study looking at influenza transmission in primary schools in Japan (5). In the latter, the authors conclude that outbreaks likely could not be supported by within-school transmission alone and suggest they can continue to exist by new infections originating from household transmission. Furthermore, in a non-pandemic period the school and household situation do not change compared to the pandemic period. The children attend the same school, the same class, with the same size, and will remain part of the same household. Therefore, our observations are also informative for the non-pandemic period, albeit that the situation outside the school and household setting differs due to pandemic control measures. This likely lowers the contribution to transmission of activities outside school and household in a pandemic period compared to a non-pandemic period.

Our observations warrant a careful interpretation. Firstly, the transmission pairs were identified as part of contact-tracing efforts and are therefore biased towards transmission events in which infector/infectee are able to identify each other. Several other locations where it is hard to identify people, such as transmission in public transport, in a shop, or on the street are therefore likely underrepresented. Secondly, the dominant variant circulating in the Netherlands in March 2021 was the alpha variant, with over 75% of cases caused by this type (6). As the infectiousness between variants change, the transmission patterns could also alter between variants. Thirdly, only those who are tested are identified as positive cases. In this period, 571,866 tests were performed among those of school age (birth year between 2003 and 2016), of which a higher number was performed among primary school children (appendix I). Therefore differences in the number of pairs between primary to secondary school compared to secondary to primary school can be due to a difference in testing among secondary school children. Fourthly, the observed transmission patterns are affected by interventions. Due to active testing and tracing, with quarantining of contacts, and extra measures in secondary schools, such as indoor wearing of masks and distance learning, as well as the reduced contact between children outside school hours, the identified transmission pairs are likely an under-representation of transmission that would have happened in a situation without measures, or with different measures. Also, because schools were highly encouraged to limit the number of contacts, the fraction of transmission between classes at school might be reduced. In addition, because secondary schools were only open for at least one day per week, whereas primary schools were open full-time for in-class education, the relative contribution of transmission for primary and secondary schoolers could change. More recent data (appendix II), from September 6^th^ until October 15^th^ 2021 for infectors and October 22^nd^ for infectees, shows very similar results, however with a lower ratio of secondary-secondary school pairs with respect to primary-primary school pairs (660:1746) compared to the data over which we report (1157:2105) despite that in this period both primary and secondary schools were fully open. Comparison between the two time periods however is difficult, vaccination of secondary school children (12-17 year olds) started on June 2^nd^ 2021, which leads to a reduced transmission in secondary school children. In addition, in the more recent period much fewer infectors were identified (27.5% vs. 39.2%) leading perhaps to a more biased sample. Fifth, no data were available on which specific school or class the infected person attended. Therefore, only transmission between primary and secondary school could be identified, rather than transmission between primary schools or between secondary schools. Exact home address of infectors and infectees was unavailable as well. However, persons in a transmission pair with the same full 6-digit postcode are in most (97.3%) cases registered on the same home address (7). Further, in a large fraction of our reported transmission pairs with setting home, the 6-digit postcode is equal for infector and infectee (89.5%). Sixth, secondary education is tiered in the Netherlands, with different schools and different number of years for the different levels of education. Only the highest level of secondary education has six years. Therefore, class S5 and S6 are for some the first years of follow-up education, the first years of working life, or indeed the last two years of secondary education. This is likely the explanation for the increased contribution of “other” in this older age range. Moreover, exam students of secondary schools (some students in S4 and S5 and all students in S6) had fully physical education whereas other secondary school classes attended school at least once per week. The interpretation of these exam years is therefore different compared to classes in primary school or class S1-3.

Which lessons are there to be learned for the rest of the pandemic? Omicron has become the dominant strain and in the Netherlands all those aged 12+ are targeted by vaccination. In-class education in secondary schools is now offered full-time, extracurricular activities have resumed, and testing criteria and quarantine guidelines are now more narrowly defined.

The data show that even with the alpha-variant, the oldest classes of primary school did participate in transmission to and from others in primary and secondary school. Given that primary schools enable transmission towards secondary schools, it might be that widespread outbreaks can be facilitated by the oldest classes in primary school, and the youngest classes of secondary education. Vaccination of children who are in higher classes of primary schools who have siblings in secondary school could therefore curb these outbreaks. We suspect that the observed lower transmission from secondary school towards primary schools is likely due to the reduced in-class learning in secondary schools at that time. Therefore, when in-class teaching is resumed, more transmission from secondary to primary schools is expected. Considering this, vaccination of children in the younger age groups in secondary education (12- and 13-year-olds) with siblings in primary school could also limit outbreaks. Hesitancy for vaccination by the Dutch National Immunisation Programme is clustered in certain identity-based schools in the Netherlands (orthodox protestant denomination and Steiner schools). The probability of large outbreaks remains in clusters of primary and secondary schools linked to these identities, even when vaccine uptake increases nationally.

In conclusion, the results provide support of transmission on a bipartite school-household network, in which households play a key role in transmission between school years and between primary and secondary schools.

## Data Availability

All data produced in the present study are available upon reasonable request to the authors

## Acknowledgements

We would like to thank for all those involved at the Municipal Health Service for their continuous effort to curtail the transmission in this pandemic, and to provide key information in the administrative system. Furthermore, we thank Marc Meurs of DUO for his input.

## Members of the RIVM COVID-19 surveillance and epidemiology team

Agnetha Hofhuis, Amber Maxwell, Annabel Niessen, Anne Teirlinck, Anne-Wil Valk, Birgit van Benthem, Brechje de Gier, Bronke Boudewijns, Carolien Verstraten, Claudia Laarman, Danytza Berry, Daphne van Wees, Dimphey van Meijeren, Don Klinkenberg, Eric Vos, Eveline Geubbels, Femke Jongenotter, Fleur Petit, Frederika Dijkstra, Gert Broekhaar, Guido Willekens, Hester de Melker, Irene Veldhuijzen, Jan Polman, Jan van de Kassteele, Janneke Heijne, Janneke van Heereveld, Jeanet Kemmeren, Kirsten Bulsink, Kylie Ainslie, Lieke Wielders, Liselotte van Asten, Liz Jenniskens, Loes Soetens, Maarten Mulder, Maarten Schipper, Marit de Lange, Marit Middeldorp, Marjolein Kooijman, Miek de Dreu, Mirjam Knol, Naomi Smorenburg, Nienke Neppelenbroek, Patrick van den Berg, Pieter de Boer, Priscila de Oliveira Bressane Lima, Rianne van Gageldonk-Lafeber, Sara Wijburg, Scott McDonald, Shahabeh Abbas Zadeh, Siméon de Bruijn, Sjoerd Wierenga, Susan Hahne, Susan Lanooij, Susan van den Hof, Sylvia Keijser, Tara Smit, Thomas Dalhuisen, Timor Faber, Tjarda Boere

## Appendix I

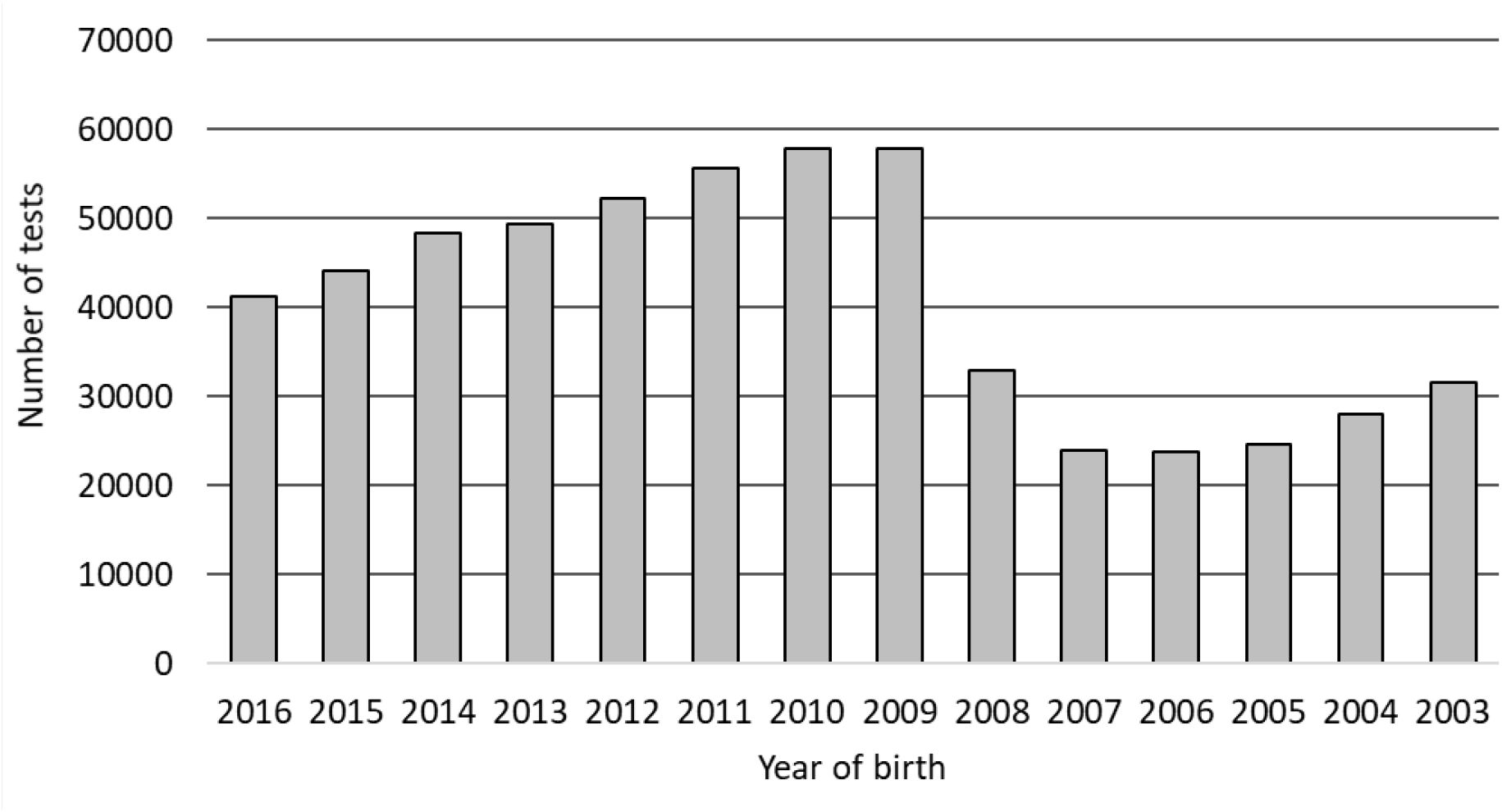
The number of tests stratified by birth cohort, over the study period, which shows that more tests were performed among primary school children compared to secondary school children.

## Appendix II

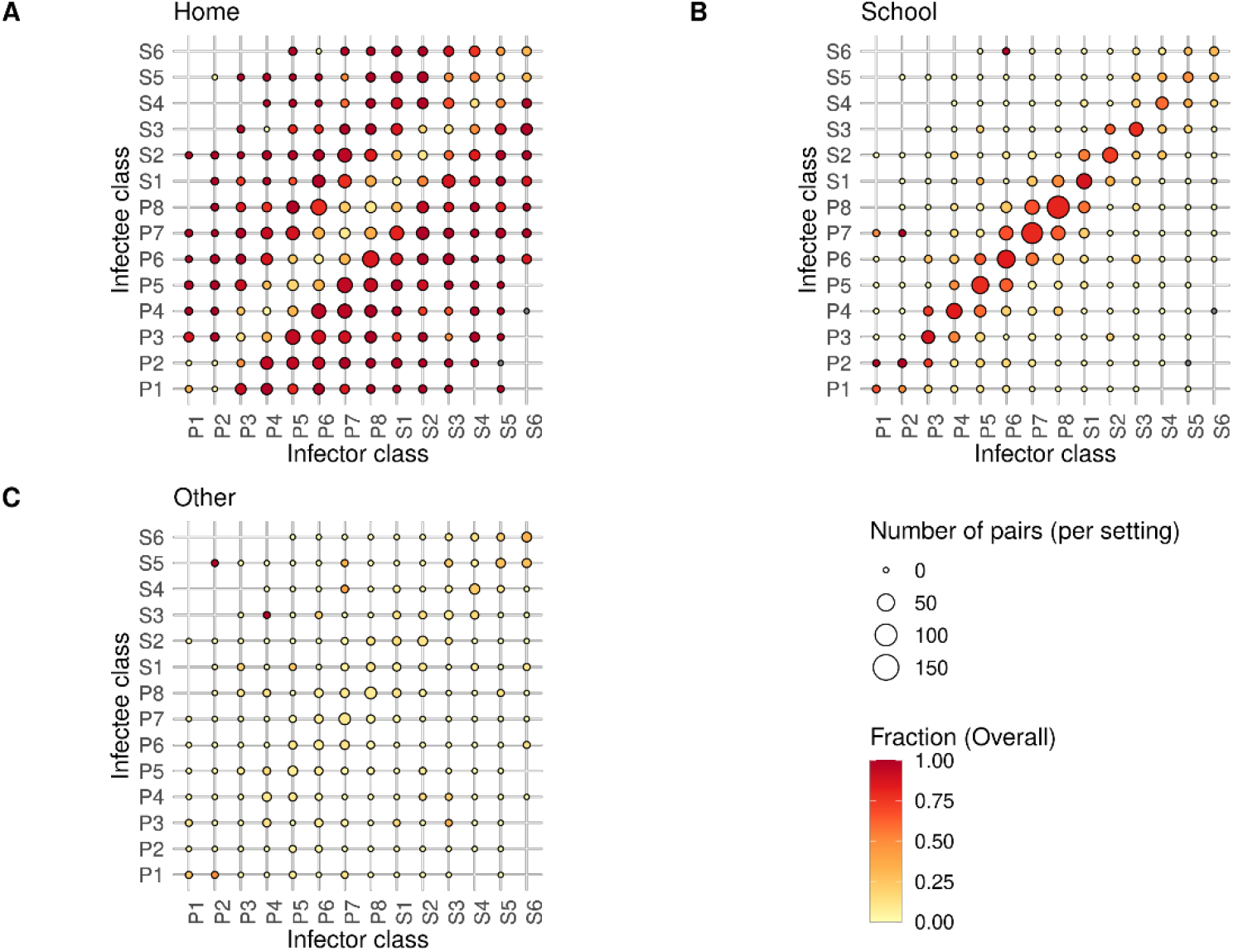
Transmission pairs by class (primary school P1-P8, secondary school S1-S6) in the period between the 6^th^ of September (start of the new school year) and the 15th of October (start of the first school holiday) 2021 for infectors and between the 6^th^ of September and the 22th of October 2021 for infectees. The size of the bubbles shows the number of pairs per setting. Transmission pairs with an unknown setting are excluded. The color of the bubbles indicates the fraction overall of transmission in **A -** the home setting. **B -** the school setting. **C -** other settings than household and school setting.

